# Educational differentials in domain specific physical activity by ethnicity, age, and gender: findings from over 44,000 participants in The UK Household Longitudinal Study (2013-2015)

**DOI:** 10.1101/19002105

**Authors:** Meg E Fluharty, Snehal M Pinto Perira, Michaela Benzeval, Mark Hamer, Barbara Jefferis, Lucy Griffiths, Rachel Cooper, David Bann

## Abstract

**Background:** The prevalence of overall physical inactivity remains high, particularly amongst socioeconomically disadvantaged groups. It is unclear however if such inequalities vary systematically by age, sex, or ethnicity, and if there are differing effects across physical activity (PA) domains.

**Methods:** We used data from a nationally representative survey of the UK, Understanding Society, with information on educational attainment (our indicator of socioeconomic position), PA and demographics collected in 2013-2015 (N= 44,903). Logistic regression analyses were conducted to test associations of education with three different PA domains (active travel, occupational and leisure time). To examine modification of the associations between education and physical activity in each domain by sex, age and ethnicity, we tested two-way interaction terms (education x ethnicity; education x sex; education x age).

**Results:** Lower educational attainment was associated with higher active transportation and occupational physical activity, but lower weekly leisure-time activity. These associations were modified by sex, ethnicity, and age. For example, education-related differences in active travel were larger for females (difference in predicted probability of activity between highest and lowest educational groups: −10% in females, (95% CI: −11.9, 7.9) −3% in males (−4.8, −0.4). The education-related differences in occupational activity were larger among males −35% (−36.9, −32.4) than females −17% (−19.4, −15.0). Finally, education related differences in moderate to vigorous leisure time activity varied most substantially by ethnicity; for example, differences were 17% (16.2, 18.7) for White individuals compared with 6% (0.6, 11.6) for Black individuals.

**Conclusions:** Educational differences in PA vary by domain, and are modified by age, sex, and ethnicity. A better understanding of physically inactive sub-groups may aid development of tailored interventions to increase activity levels and reduce health inequalities.

## Introduction

Physical activity is an important modifiable determinant of health [1]. In particular, leisure time physical activity’s (LTPA) benefits are well documented including improvements in the musculoskeletal system, maintenance of healthy weight, protection against cardiovascular disease, and reduction in depression and anxiety symptoms [1, 2]. However, there is a global trend towards high levels of leisure time physical inactivity which is estimated to contribute to ∼6-10% of major non-communicable diseases, ∼5.3 million deaths annually [3-5], and ∼$67.5 billion per year in health care expenditure [6].

Physical activity can be accrued through multiple domains (e.g. active transportation, leisure time, occupation, and housework/DIY), which may have differing impacts on health outcomes [7, 8]. For example, leisure-time physical activity (LTPA) is thought to be beneficial to physical health and wellbeing, while labour-intense occupations may increase risk of musculoskeletal strain [9, 10]. Therefore, examining these different domains may provide evidence to help inform where possible interventions could targeted. Understanding what is driving differences in activity participation overall, as well as in different domains of physical activity, may also help to identify which forms of activity could be intervened on to reduce socioeconomic disparities in health.

Recent systematic reviews find evidence of socioeconomic disparities in LTPA in high-income countries [11, 12] that have persisted across recent decades [7, 13]. Additionally, lower education was associated with higher risk of future declines in LTPA [14]. Alongside indicators of socioeconomic position, a number of other sociodemographic factors, including ethnicity, gender, and age are associated with physical activity [7, 13, 15]. Different levels of participation are reported across ethnic groups in the UK, with those of ‘mixed’ ethnicity having the highest prevalence of activity [16-18], and South Asians the lowest [18, 19]. Numerous factors may explain these differences in participation including personal, socioeconomic, cultural, and environmental factors [16-19]. Additionally, overall global physical activity levels are lower for women and older adults [20, 21]. These differences may result from gender-specific events over the life course (e.g. during pregnancy) impacting women’s physical activity to a greater degree than men’s, or suggest differences in physical activity opportunities available for women (safety, accessibility, or availability around caring/childcare duties) [20, 22]. Furthermore, lower physical activity levels at older ages may reflect a number of factors including changes in work and health status [18, 19, 22, 23].

Educational disparities in physical activity may arise though a number of routes including due to differences in knowledge of the health impacts of LTPA, material pathways, and potentially due to selection into neighbourhoods which differ in the suitability for outdoor physical activity [24, 25]. However, educational inequalities in physical activity may be modified by ethnicity, age, and sex [8, 26]. For example, low-level education occupations are more physically demanding among men [8, 27]; however, these individuals’ display lower engagement with leisure-time activities [28]. Moreover, evidence from the US has indicated education-related disparities across domains [8]. However, these associations have not yet been investigated within the UK. Previous studies that have investigated associations of different indicators of socioeconomic position including education and demographics with physical activity outcomes are limited by only investigating one specific domain [29-34], or use population samples from specific regions within the UK [35, 36]. Thus, important gaps remain in our understanding of the nature of inequalities in physical activity outcomes. These are important to fill given their purported mediating role in socioeconomic inequalities in key health outcomes such as premature mortality [37].

We sought to address the above-mentioned gaps in the literature by investigating educational disparities in physical activity across travel, leisure, and occupational domains. Additionally, we examine if the associations between education and domain specific physical activity are modified by ethnicity, age, and sex. We hypothesised that the associations between education and domain specific physical activity will be modified by ethnicity, age, and sex. Additionally, we hypothesised that lower education status will be associated with lower physical activity during leisure time, but higher activity in active travel and work. A large household panel study was used (Understanding Society), which benefits from national representation, oversampling of ethnic minority groups, and detailed measures of physical activity across leisure, travel, and work domains.

## Methods

### Participants

Understanding Society: the UK Household Longitudinal Study (UKHLS) is a nationally representative study which started in 2009 aiming to recruit individuals in 40,000 households [38]. The study annually samples all individuals in the household over the age of 10. Additionally, sample members are followed when they leave the household, and new individuals join the study as they become part of an existing study member’s household. Information is collected from participants on a range of information including wellbeing, health, home, family and employment. Detailed study information and sampling methodology can be found elsewhere [38]. All participants consent for use of their anonymised survey information, and data for this study were accessed through the UK Data Service (https://www.ukdataservice.ac.uk/).

The sample for our analysis includes adult (20 years or over) responders who took part in Wave 5 (2013-2015) and responded to demographic and physical activity questions via interviewer led and self-completed questionnaire. Wave 5 was chosen as this was the most recent wave of data collection including physical activity questions.

### Measures

#### Domain specific physical activity

Active travel was measured in currently employed individuals and those not working from home via the question ‘*how do you usually get to your place of work?’* Responses were collapsed into a binary variable of ‘non-active’ (car, bus, or train/metro) or ‘active’ (walking or cycling). Occupational physical activity was measured by asking participants whether their job was mainly physical or not (categorised as ‘not physical’ and ‘physical’). Finally, leisure-time physical variables were created from participant responses to the ‘Taking part Survey’ [39] which indicated how often they participated in a series of prelisted sports and activities. Sports were then grouped into two categories based on their average metabolic equivalent of task (MET), those with METs of ≥3 were categorised as moderate-to-vigorous and METs 1.5-2.9 as light [40]. Frequency of participation in each MET-group was categorised as weekly or non-weekly.

#### Socio demographics

Highest educational attainment was self-reported and categorised into three groups: ‘degree or higher, ‘school diploma/other qualification’ (e.g. A levels and vocational diplomas) and ‘GCSEs and below’. Ethnicity was self-reported and responses were collapsed into ‘White’, ‘Black’, ‘Asian’ and ‘other ethnicity.’ These broad ethnic groupings include minority groups (e.g. ‘White’ includes all white minorities such as Irish and Polish, Black includes Black-African and Black-Caribbean, and those of smaller sample sizes such as Arab and mixed-ethnicity were included in ‘other’). Age at the time of interview was categorised into ten year age groups (from ages 20-60). Older adults were grouped from >60 years, and those below 20 were excluded from the analysis to ensure comparable sample sizes in the higher education groups—alternative groupings did not substantially affect the results (data available upon request).

#### Statistical analysis

We first cross-tabulated educational attainment by age, sex, and ethnicity. Next, logistic regression analyses were conducted to examine associations of education, sex, age, and ethnicity with physical activity in each domain. Analyses were assessed before and after mutual adjustment for each demographic variable. Those with missing demographic and education data yet valid outcome data were excluded from analysis (N= 1,642), similarly those below the age of 20 were excluded (N= 3, 050); analytical samples for travel, occupational, and leisure were N= 18,404, N= 22,287 and N= 40,270 respectively. A flow diagram (Supplementary Figure S1) displays the final sample size for each outcome. Sample sizes varied with each outcome due to specific questionnaire routing (e.g. only employed individuals who commuted to work were asked about active travel). Complete case analysis (sample restricted to those with valid demographic data and one domain of physical activity) was undertaken as sample sizes varied across outcomes due to specific exclusion criteria per question [41]. Finally, to examine possible effect modification of the associations between education and physical activity in each domain, we included two-way interaction terms (education x ethnicity; education x sex; education x age) in addition to the relevant first order terms in the same model. Analyses were weighted according to sample design and attrition [38]. All Analyses were conducted in Stata, version 15.0 (StataCorp LP, College Station, Texas).

## Results

All demographics (ethnicity, sex, and age) were independently associated with educational attainment and with physical activity in each domain (p< 0.001; see Table 1 and Supplementary Table S1). Lower educational attainment was associated with higher active transportation and occupational physical activity, but lower weekly light and moderate leisure-time activity.

**Table 1.** Ethnicity, sex, age and domain-specific physical activity by highest educational attainment in Understanding Society (2013-2015)

| Demographics | Highest educational attainment |  |  |  |  |  |
| --- | --- | --- | --- | --- | --- | --- |
|  | GCSEs and lower |  | School diploma<br>& other<br>qualifications |  | Degree/higher |  |
|  | N | (%) | N | (%) | N | (%) |
| **Ethnicity** |  |  |  |  |  |  |
| White | 11638 | 33.7 | 10424 | 30.2 | 12424 | 35.9 |
| Black | 363 | 25.1 | 400 | 27.6 | 685 | 47.3 |
| Asian | 1106 | 32.2 | 877 | 25.5 | 1445 | 42.1 |
| Other ethnicity | 201 | 22.1 | 235 | 25.9 | 472 | 51.9 |
| Missing | 538 | 36.1 | 491 | 33 | 401 | 26.9 |
| *P value* | <0.001 |  |  |  |  |  |
| **Sex** |  |  |  |  |  |  |
| Male | 5888 | 30.5 | 6388 | 33.1 | 6927 | 35.9 |
| Female | 7958 | 35.3 | 6039 | 26.8 | 8500 | 37.7 |
| *P value* | <0.001 |  |  |  |  |  |
| **Age** |  |  |  |  |  |  |
| 20-29 years | 1462 | 23.9 | 2363 | 38.6 | 2271 | 37.1 |
| 30-39 years | 1653 | 24.3 | 1776 | 26.1 | 3345 | 49.2 |
| 40-49 years | 2433 | 29.1 | 2339 | 28 | 3570 | 42.7 |
| 50-59 years | 2455 | 32.6 | 2221 | 29.5 | 2822 | 37.5 |
| 60+ | 5843 | 44.8 | 3728 | 28.6 | 3419 | 26.2 |
| *P value* | <0.001 |  |  |  |  |  |

*Participants from wave 5 (2013-2015) of Understanding Society with data on educational attainment, demographics, and physical activity*
*P value = chi<sup>2</sup> Missing = missing data; where not shown there were no missing data.*

### Active travel to work

Active travel was lowest amongst highly-educated individuals, older ages, and males; there was little evidence for associations with ethnicity (See Tables 2). The magnitude of education-related disparities were largest among females (education x sex P<0.001) and Black individuals (education x ethnicity P= 0.038) (See Figure 1 & Supplementary Tables S4-6). For example, the estimated difference in the probability of using active transportation in the highest versus the lowest educational group was −10% (95% CI: −11.9, 7.9) amongst females and −3% (−4.8, −0.4) among males. Results for this domain, and all others, were similar when restricting to those with valid demographic and physical activity data, or when not making this restriction (Supplementary tables S2-S3).

**Table 2.**
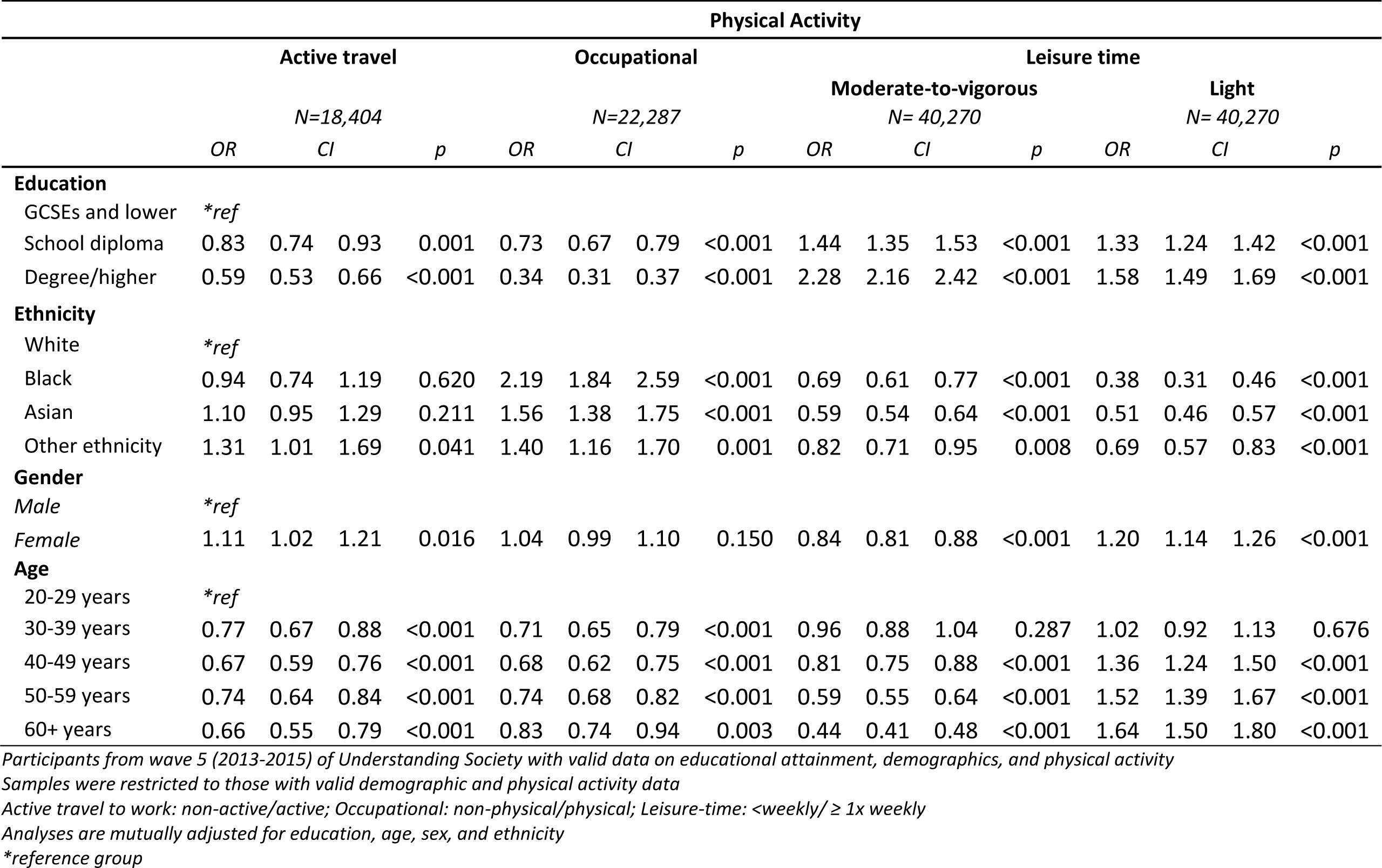
Mutually adjusted associations between educational attainment, demographics, and domain specific physical activity outcomes, mutually adjusted for all education and demographic factors.

**Figure 1.**
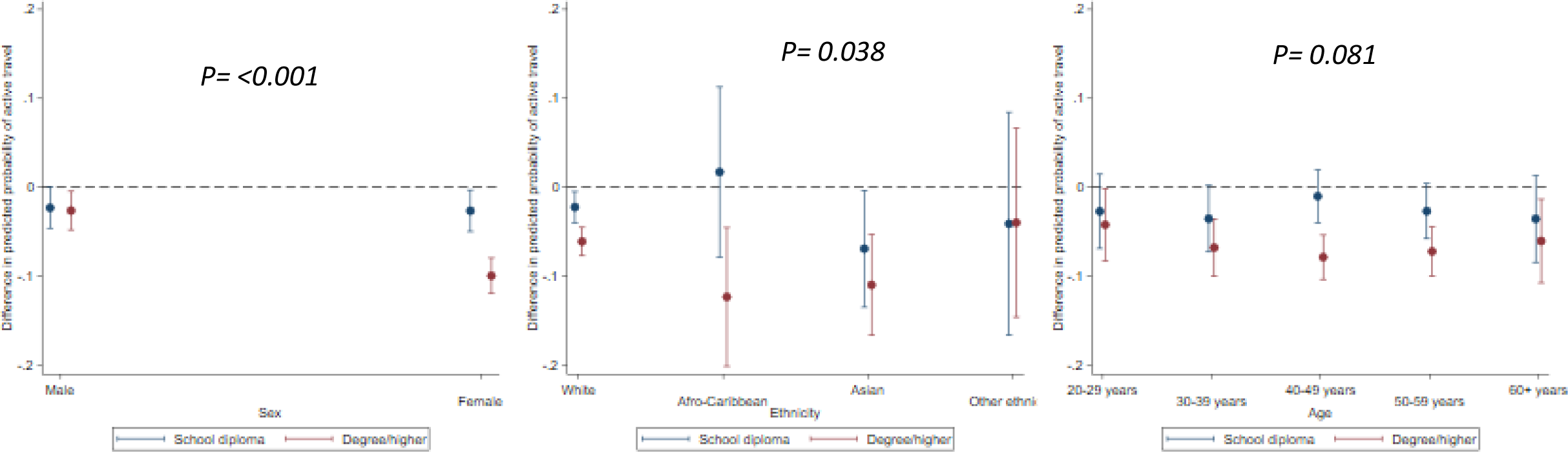
Educational differences in active travel by sex, ethnicity, and age.

### Occupational activity

Physically active occupations were lowest amongst highly-educated individuals, White individuals, and those aged over 20-29; there was no evidence for association with sex (see Tables 2). The magnitude of education-related disparities were largest among men (education x sex P <0.001), and those aged 30-39 (education x age P= 0.001) (See Figure 2 and Supplementary Tables S4-6). For example, the estimated difference in the probability of a physical occupation in the highest versus lowest educational group was −35% (−36.9, −32.4) for males and −17% (−19.4, −15.0) for females.

**Figure 2.**
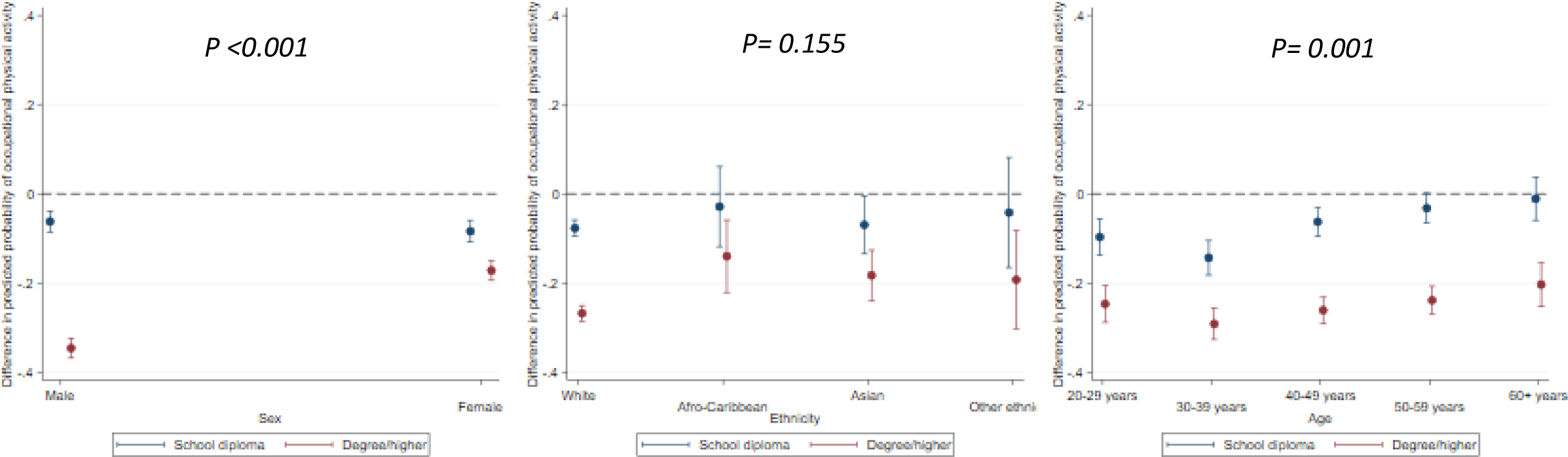
Educational differences in occupational physical activity by sex, ethnicity, and age.

### Moderate-to-vigorous and light Leisure time activity

Weekly moderate-to-vigorous LTPA was highest amongst highly educated individuals, Whites, males, and younger adults. While weekly light leisure time activity was highest in highly-educated individuals, Whites, females, and older adults, and lowest in Black individuals (See Tables 2-3).

The magnitude of education-related disparities in weekly moderate-to-vigorous leisure time activity was largest among White and Asians individuals (education x ethnicity P= 0.001), and those aged 40-49 and 50-59 (education x age P= 0.008) (See Figures 3-4 and Supplementary Tables S4-6). For example, the estimated probability of weekly moderate-to-vigorous leisure time activity in the highest versus lowest educational group was 17% (16.2, 18.7) for white individuals, compared with 6% (0.6, 11.6) for Black individuals, 16% (12.8, 19.1) for Asian individuals, and 13% (6.0, 19.5) for those of other ethnicity.

**Figure 3.**
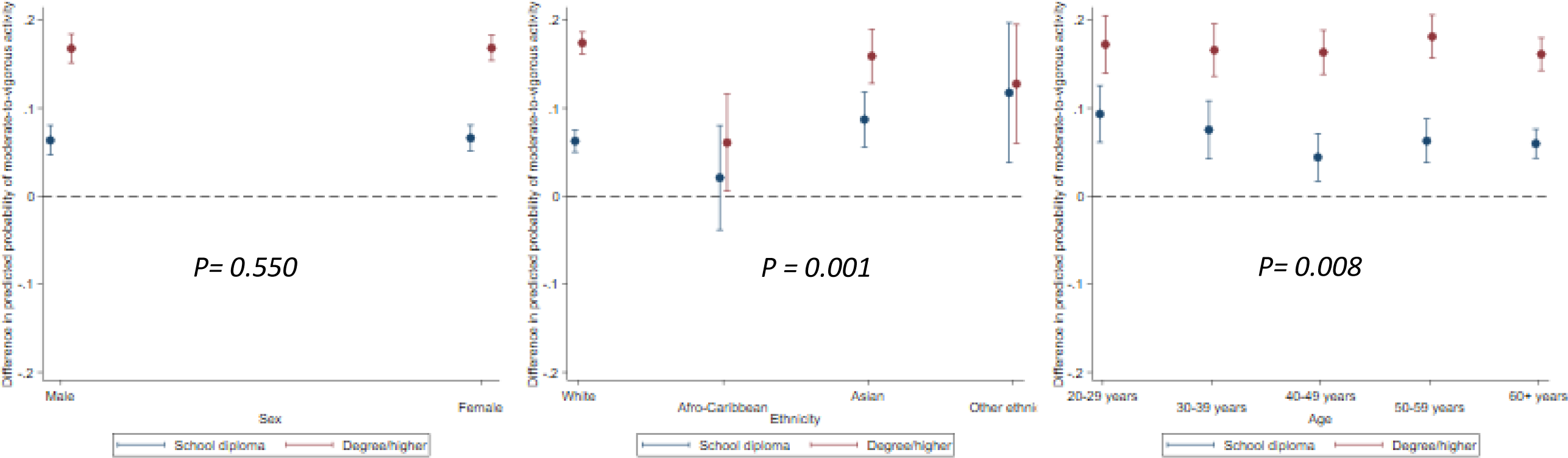
Educational differences in moderate to vigorous intesity activities by sex, ethnicity, and age.

**Figure 4.**
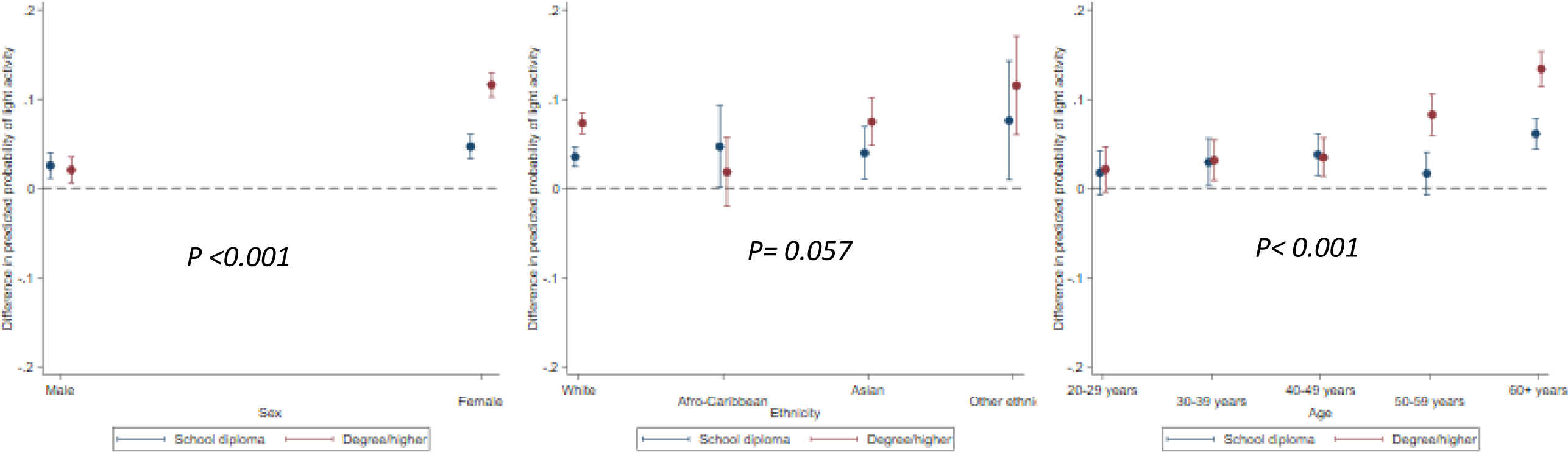
Educational differences in light intesity activities by sex, ethnicity, and age.

The magnitude of education-related disparities in weekly light leisure time activity was largest among females (education x sex P< 0.001) and individuals aged 60+ (education x age P< 0.001); there was little evidence for associations with ethnicity (See Figures 3-4 and Supplementary Tables S4-6). For example, the estimated probability of weekly light leisure time activity in the highest versus lowest educational group was 13% (11.4, 15.3) for those aged 60+ compared with 8% (5.9, 10.6) for those ages 50-59, 3% (1.3, 5.6) for those 40-49, 3% (0.9, 5.4) for those 30-39, and 2% (−.04, 4.7) for those aged 20-29.

## Discussion

### Main findings and interpretations

In a large nationally representative dataset educational attainment was associated with physical activity across three key domains; individuals with higher education were less likely to engage in active travel and occupational physical activity, but more likely to engage in leisure-time activity. These associations were modified by ethnicity, age, and sex. For active travel disparities were largest among females and Black individuals. For occupational physical activity disparities were largest among men and those aged 30-39. For moderate-to-vigorous leisure time disparities were largest among White and Asian individuals and those aged 40-49 and 50-59. Finally, for light leisure time disparities were largest among females and those aged 60+.

Previous evidence suggests a number of possible disparities such as health status [42], environment [43, 44], cultural preferences [17, 45], financial resources [37, 46, 47], perceived safety [32, 48], and domestic requirements [20, 22] may exist within each sociodemographic group of the same levels of educational attainment. These differences may in turn explain the observed modification of education disparities in physical activity. These potential explanations require future empirical investigation.

### Strengths and Limitations

Strengths of this study include a large nationally representative sample, enabling us to identify the previously seldom-examined role of ethnicity as a modifier of the relationship between educational attainment and physical activity across different domains. We also examined activity outcomes across three domains; previous studies investigating associations of physical activity typically use a single physical activity outcome measure, capturing either a ‘leisure’ or ‘unspecified’ variable [7, 8].

There are also a number of limitations to consider. First, while we obtained information across multiple domains, we lack detailed information on activity duration. Nor did we consider perceptions of the local environment including safety which may affect physical activity [32, 48]. However, the leisure time activity measures used followed expected patterns for these leisure time categories by sex and age [49, 50]. Second, we did not capture physical housework as a domain, which includes domestic and cleaning tasks, gardening, and DIY [51]. Third, all physical-activity measures were captured via self-report; while this is needed to investigate domain-specific evidence, it may be subject to recall bias with individuals’ either over or under reporting their levels of physical activity [52, 53]. For example, previous evidence has shown that males and those who achieved fewer years in education were more likely to overestimate their physical activity levels than females and those with greater levels of education, respectively [54]. Additionally, measurement error in self-reported physical activity may account for differential reporting bias across cultures and social strata [55, 56]. Fourth, active travel only included working adults in the analyses of the travel and occupational domains, which are only relevant to those in-work; therefore, investigation of multiple types of physical activity among retirees or those unable to work warrants consideration in the future. Fifth, bias may be introduced through excluding missing data and non-responders, although missing data due to item missingness (as opposed to specific question gating) was low and therefore bias unlikely. Sixth, due to the cross-sectional design we cannot separate out age from birth cohort, and future cross-cohort studies are therefore required to address this.

Finally, this study identified cross-sectional associations of education with physical activity across domains, as well as education-related differences in ethnicity, sex, and age in the physical activity domains. However, associations of educational-attainment with physical activity may be partially explained by reverse causality [57], particularly at younger ages. Further longitudinal analyses are required to determine the direction of association and additionally identify the mediators of the disparities observed.

### Implications for practice, and policy

Our findings may have important implications for practice and policy. The inequalities in leisure time physical activity observed—across both light and moderate-vigorous activity—suggests that policies are required to reduce these inequalities given the multiple anticipated effects on health. Population-level or targeted interventions may be used to reduce the sizeable modification across demographic sub-groups. For example, there was a 13% difference in probability of engaging in light LTPA in those aged 60+ compared to 2-3% of those aged 20-39, suggesting lower educated older adults would benefit most from interventions regarding this domain of physical activity. Furthermore, in line with previous evidence [27] we also found that those of lower educational attainment were more likely to possess physically demanding occupations. This difference is important, if the health consequences of occupational physical activity are less favourable or detrimental compared with LTPA [9, 10]. Efforts to increase leisure time activity and its inequality should consider co-occurring differences in occupational activity. Finally, lower engagement in domain-specific physical activity were also found in those with high levels of education attainment. For example, there was a 10% difference in active travel among high to low educated women compared with 3% in men. Previous evidence has found similar sex differences in cycling to work; however, similar rates of men and women for leisure-time cycling [58]. Suggested means of increasing active travel include the provision of safe walking and cycling travel routes, accessible bike locks, and changing facilities.

Finally, our findings may have implications for future studies which investigate inequalities in physical activity outcomes. Existing studies typically adjust for the sociodemographic factors we investigated as potential modifiers. Given the evidence for modification that we found, such analyses may provide biased estimates of the magnitude of inequalities that operate in particular population sub-groups

## Conclusions

In summary, we found sex, age, and ethnicity modified associations between educational attainment and multiple physical activity outcomes. Our findings imply there may be unequal access or additional barriers to physical activity across both education and demographic sub groups. Better understanding the characteristics of physically inactive sub-groups may aid development of tailored interventions to increase activity levels and reduce health inequalities.

## Data Availability

Data for this study were accessed through the UK Data Service.

https://www.ukdataservice.ac.uk

## Acknowledgements

This project is part of a collaborative research programme entitled ‘Cohorts and Longitudinal Studies Enhancement Resources’ (CLOSER) funded by the ESRC (http://www.esrc.ac.uk) (ES/K000357/1). DB and MF are also supported by the Academy of Medical Sciences/the Wellcome Trust “Springboard Health of the public in 2040” Award (HOP001\1025). SPP is funded by a UK Medical Research Council Career Development Award (MR/P020372/1).

Understanding Society is an initiative funded by the Economic and Social Research Council and various Government Departments, with scientific leadership by the [Institute for Social and Economic Research](http://www.iser.essex.ac.uk/), University of Essex, and survey delivery by [NatCen Social Research](http://natcen.ac.uk/) and [Kantar Public](http://www.kantar.com). The research data are distributed by the UK Data Service. MB is funded by ESRC ES/N00812X/1.

## Notes

### Competing Interest Statement

The authors have declared no competing interest.

### Author Declarations

All relevant ethical guidelines have been followed and any necessary IRB and/or ethics committee approvals have been obtained.

Any clinical trials involved have been registered with an ICMJE-approved registry such as ClinicalTrials.gov and the trial ID is included in the manuscript.

